# Management of Intraductal Papilloma Using Vacuum Assisted Breast Surgery (VABB/VAE):- A Single Institute Study

**DOI:** 10.1101/2024.01.23.24301707

**Authors:** Rohan Khandelwal, Pooja Sharma, Dev Desai, Sambit Mohanty

## Abstract

Intraductal Papilloma has a higher upgradation rate despite being a benign tumor and warrants an immediate excision. Surgical removal is the gold standard but carries poor cosmetic results and requires general anesthesia. Ultrasound-guided Vacuum Assisted Excision (US-VAE) was used to treat 47 patients after due ethical permissions. 32 patients went US-VAE under local anesthesia and all patients were happy with the cosmetic outcome. The excision was done as a daycare procedure in all patients except indicated by the Anesthetist. Requirements of post-procedure analgesia were also minimal and only 3 patients required prolonged analgesia. The advantages of VABB-guided surgery are faster recovery, scarless procedure, and local anesthesia, thus making it an ideal OPD and daycare procedure for Intraductal Papilloma Excision.

**Introduction:** Intraductal papilloma, although benign, has a high upgradation rate to malignant breast cancer. Surgical excision is necessary, but the rate of complications is high. Ultrasound-guided vacuum-assisted excision (US-VAE) is a new technique derived from the VABB biopsy method that can give better cosmetic results as well as fewer surgical complications.

**Objectives:** Understanding the utility of US-VAE in the diagnosis and treatment of intraductal papillomas of the breast.

**Methodology:** With permission from the Ethics Board and consent of the patients, records of 47 patients were recruited to undergo the US-VAE procedure. USG was used to stage the tumor with Mammography and CNB as needed and patients with single duct or adjacent duct lesions were included while the rest, including patients with malignant changes, were excluded. The Bard VABB biopsy machine and the 7G probe were used by highly trained breast surgeons to perform the procedure.

**Results:** Of the 47 patients included, 32 patients under local anesthesia were operated while others were under general anesthesia according to the preference of the patient and the level of apprehension. The patients received oral paracetamol as the only painkiller for 5 days. Only 3 patients required prolonged analgesics. 7 patients had prolonged bruising (lasting more than one week), which was the most common complication. All patients (100%) were happy with the cosmetic result after the procedure.

**Conclusions:** The advantages of VABB-guided surgery are faster recovery, scarless procedure, and the ability to perform the surgery under local anesthesia, making it an OPD and daycare procedure.

## Introduction

Intraductal breast papillomas are common benign tumors of the breast that arise from the ductal tissue. They often present with bloody/severe nipple discharge from a single duct and are typically found between the ages of 30 and 55. They can be visualized by ultrasound, and in equivocal cases, an MRI of the breast is required. As these lesions tend to be associated with papillary ductal carcinoma in situ, they require surgical excision following a preoperative core biopsy. (1) Histopathologically, it is a benign growth of the breast ducts in which the duct epithelial cells proliferate abnormally. Risk factors include contraceptive use, hormonal replacement therapy, lifetime estrogen exposure, and a positive family history. (2)

Although benign, intraductal papilloma can undergo surgical transformation to atypical ductal hyperplasia, DCIS, or carcinoma. Approximately a 10% upgrade rate is found with Core Needle biopsy (3). As this possibility of upgrading is present at the time of excision, therapeutic management involves surgical excision and complete removal of the tumor. (1) Close observation or open excision ( microdochectomy) are the usual treatments for benign breast lesions. Open excision in the form of microdochectomy involves the removal of the duct lesion along with the duct, and this usually involves a periareolar incision and excessive scarring. (4)

VABB (Vacuum Assisted Breast Biopsy) is a procedure used for biopsy in patients with breast tumors. It is a minimally invasive technique. In some cases, the lesions were found to be so small that very little tissue remained after the biopsy was taken. It became evident that a slight modification in the technique could be used for the excision of this tissue. From this, the US-VAE technique was developed. (4-6) Given the option, it is now preferred by more and more women for its satisfactory cosmetic result and few postoperative complications.

This study attempted to investigate the characteristics of the lesion and histologic factors that influence the rate of excision with vacuum-assisted core biopsy to determine the utility of ultrasound-guided vacuum-assisted core biopsy in the diagnosis and treatment of intraductal papillomas of the breast.

## Methodology

After ethical considerations and permission from the relevant IRB, a total records of 47 patients who underwent VABB-VAE (Vacuum Assisted Breast Biopsy - Vacuum Assisted Excision) for duct papilloma at the tertiary care hospital from May 2021 to January 2023 were enrolled. The procedure was carried out by trained surgeons for some time under the proper surgical conditions. These 47 patients had USG-guided VABB excision performed using a Bard VABB biopsy machine and a 7G probe. Clinical and demographic data were obtained from medical records. The patients underwent scheduled follow-up at 3 months and 6 months after surgery. There is a minimum of 6 months of follow-up period for each patient.

The selection of patients for the procedure was performed only after USG diagnosis by an expert breast radiologist. In case the USG findings were equivocal, the patients were subjected to CEMRI on the breast. Patients who have single-duct papilloma or multiple-duct papillomas in adjacent ducts were included according to the USG report. Patients with USG reports suggesting multiple duct papillomas in wide ducts or patients diagnosed with papillary DCIS according to core needle biopsy were excluded.

These patients underwent the US-VAE procedure. Types of anesthesia used, pain after the procedure, duration, analgesic molecules, intraoperative and postoperative complications, cosmetic results, and recurrence of the injury were recorded to correlate and analyze.

Software such as Excel and SPSS20 was used to analyze using tests. The Average ± Standard Deviation format was used for the major information.

## PROCEDURE

The procedure was performed as a daycare procedure or as a 24-hour admission based on the patient’s need. The procedure was performed by a breast surgeon, trained with VABB, after giving informed consent. Before the procedure, an ultrasound was performed to see the location and size of the lesion and to decide the entry of the probe. The Bard VABB biopsy machine and the 7G probe were used.

Adequate tumescent anesthesia was administered with Lignocaine with Adrenaline, Bupivacaine, and saline, on the skin and around the mass. (5)

A small incision was made with an 11-no blade. The probe was inserted into the lesion and the removal process of the tumor was visualized by ultrasound. The lesion was automatically sucked into the tubular sampling groove through negative pressure and came out piecemeal. The probe was slowly rotated to complete 360 degrees until the tumor was completely removed. Any remaining lesions were examined on an ultrasound examination after removal and the site and probe tract were compressed for 10–15 minutes to prevent bleeding. A post-procedure clip was inserted into the cavity. The incision was closed with Monocryl and compression dressing.

**Image 1:**
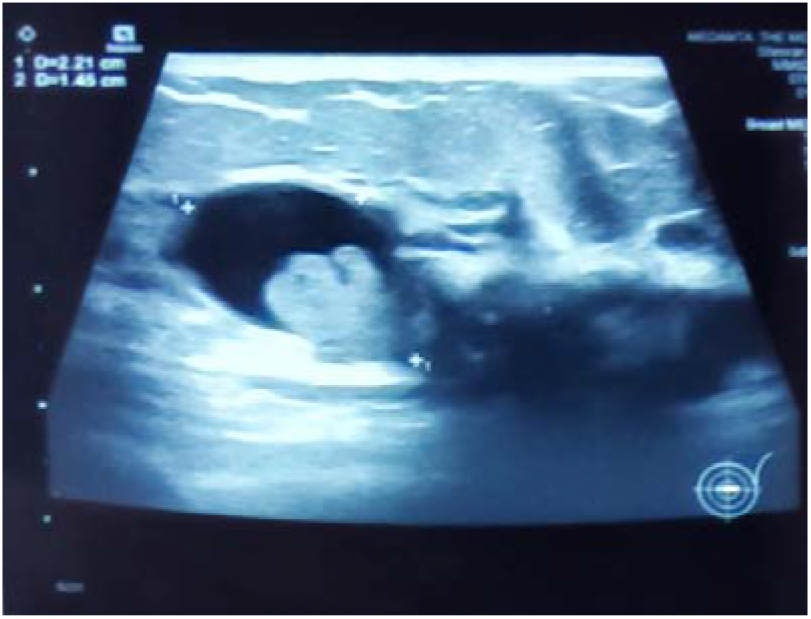
Ductal Papilloma in USG.

**Image 2:**
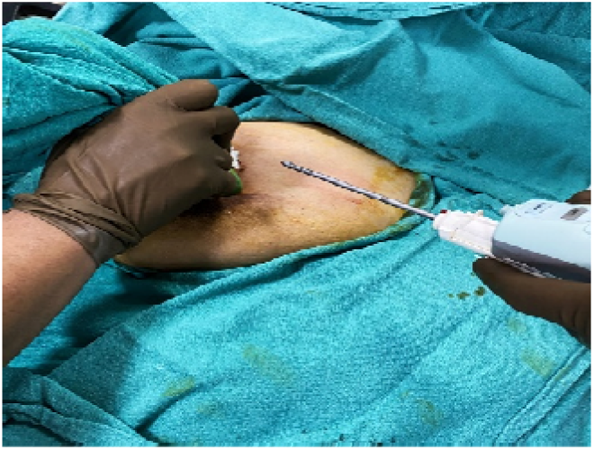
Excision using. The Bard VABB biopsy machine and 7G probe.

**Image 3:**
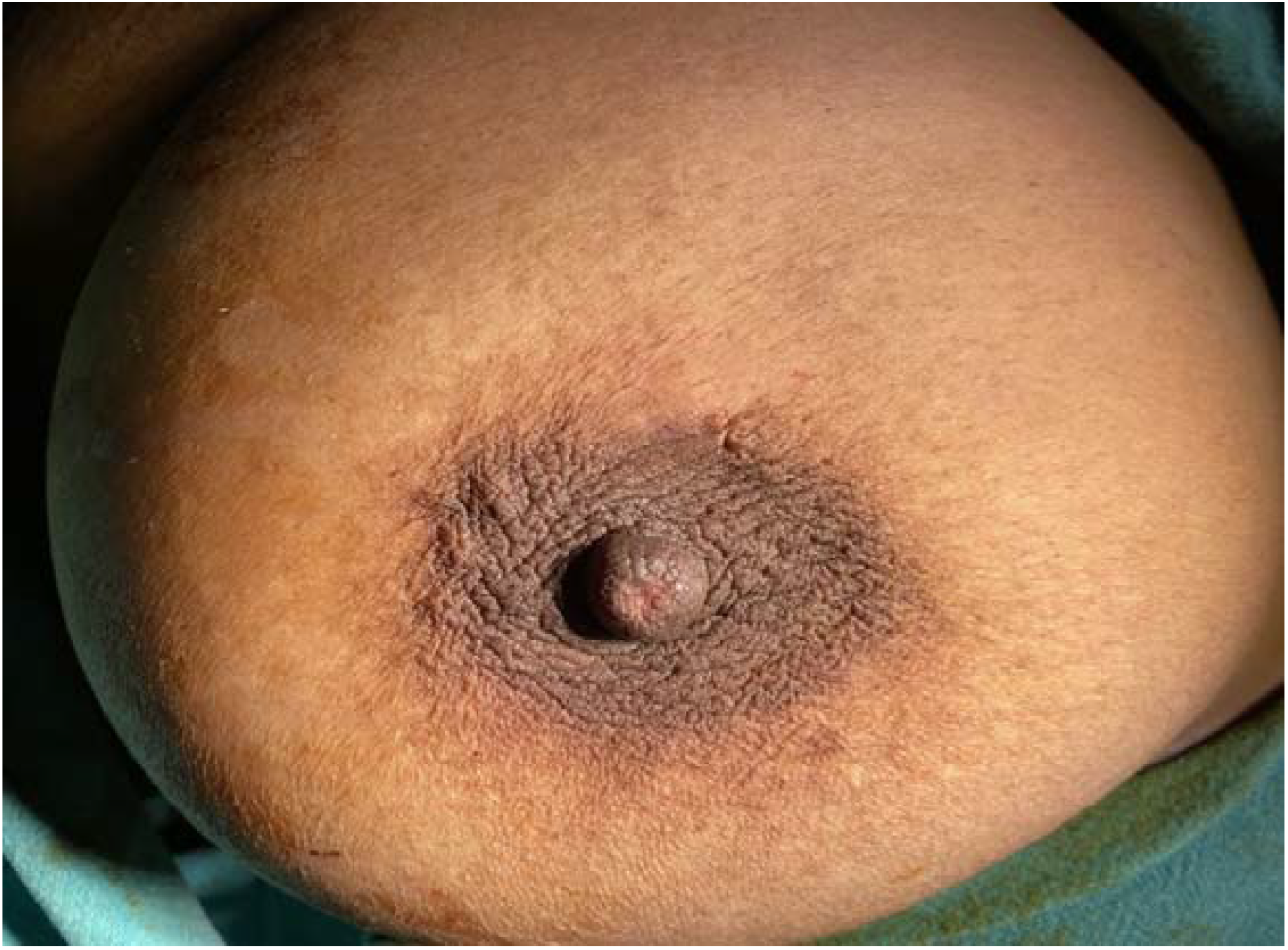
Immediate post-excision breast image showing minimal surgical wound.

## Results

This procedure was performed to see its effectiveness for the excision of noncomplicated intraductal papilloma. Therefore, all 47 patients included in the study had intraductal papilloma. All patients were women in the 30-55 age group and were treated with US-VAE as an OPD procedure with only 4 patients undergoing the procedure in the operating room with only admission a single day from morning to night. Patients who opted for general anesthesia were discharged the following day according to the Anesthetist’s requirement.

All lesions were in the 4 to 15 mm mean size range. All lesions were in the category of BIRADS 3 & 4a according to the USG reports.

The most general presentation of patients to the hospital was due to bloody nipple discharge as it was the most common complaint. Other complaints included discomfort and heaviness. Some patients also came to the hospital after finding small nodules while performing a breast self-examination.

Of the 47 cases included in the study, 32 (68.08%) patients were operated under local anesthesia, the rest 15 (31.92%) patients were under general anesthesia. The decision was based on the patient’s preference and the level of apprehension and had nothing to do with the size of the lesion. Whenever possible, the patient was given autonomy for their choice of anesthesia.

A single dose of antibiotic was administered for antibiotic coverage to prevent infection at the surgical site. No infections or discharges were reported after the procedure in any of the patients.

Post-procedure pain management was performed in patients with oral paracetamol It was the only analgesic administered to these patients until postoperative day (POD) 5. Only 3 (6.3%) patients required prolonged analgesics. In patients who needed analgesic coverage even after 5 days, a combination of Diclofenac and Paracetamol was administered. Pain on the visual analog scale used in POD-1 and POD-5 was less than 4 in 43 (91.4%) patients, while only 4 (8.6%) patients had rated it as more than 6.

Only 4 (8.5%) patients showed prolonged bruising (lasting more than one week), which was the most common complication. Other complications included mild pain, especially with movement, and discomfort. No cases of vasovagal symptoms, hematoma, or collection in the cavity were reported.

All patients (100%) were happy with the cosmetic result after the procedure. Monocryl sutures were used intradermally to approximate the incision site in all patients. Hence, cosmesis was at 100% because VAE surgery does not leave a scar behind, while the open excision leaves a scar.

No patient showed upgradation to DCIS post-surgery. No recurrence has been observed in any patient in the 6-month follow-up period according to the guidelines.

100% of the patients showed complete excision and had no recurrent lesions at the end of 6 months. No recurrence or progression to malignancy of the lesion is reported in the 47 patients and no patients are lost to follow-up. No mortality is reported in all of these patients.

## Discussion

Inra-ductal papillomas are common benign breast tumors arising from the ductal tissue. They can be upgraded to Papillary DCIS. This poses a requirement for the correct identification of the lesion and removal of it.

Mammographically occult intraductal papillomas are hard to diagnose despite the use of USG. Noninvasive diagnostic modalities such as magnetic resonance imaging can be used, as well as Ductography and Ductoscopy are invasive modalities. (7) Recently, MR-Galactography has shown diagnostic value (8)

Once confirmed, excision of the tumor is imminent. Surgical excision has remained the mainstay therapeutic modality. Newer surgical techniques have been developed and perfected to decrease the problems and complications that arise with surgical excision.

The main problem with surgical excision is the size of the incision. A larger incision for tumor removal means a deeper plane of anesthesia and a worse postoperative period. US-VAE uses a very small knick incision compared to surgical excision. (6) The benefits of the small incision here are shown by the fact that monocryl can be used to close the incision. With proper approximation and compression, a cosmetic result of 100% can be achieved. As this leaves no scar, 100% patient satisfaction can also be ensured.

A small incision would mean a shallower plane of anesthesia can be used, meaning faster postoperative recovery. It also means that a smaller amount of analgesics would be adequate for pain management in patients. (9)These features can be noticed because even after performing the procedure in most patients under local anesthesia with lignocaine, only oral paracetamol for 5 days is sufficient in most patients to manage dolor. The amount of pain is not associated with the volume of tissue excised with the formation of the hematoma or with the age of the patient. (10)

Surgical excision is also known to cause hematoma, vasovagal symptoms, and fluid collection as the cavity is closed and a blind sac is created. The occurrence of these complications is very low while using the US-VAE technique. (11)

US-VAE also showed great results with 0 recurrences at 6 months of follow-up. Intraductal papillomas are known to improve and become malignant, so regular follow-up must be carried out in these patients. (12)

## Conclusion

The US VAE procedure offers an overall advantage over surgical excision in all categories, including patient satisfaction and recurrence. The benefit of avoiding a major surgical scar outweighs the almost nil drawbacks, the main being the need for a trained surgeon for the procedure. That being said, similar to surgical excision, regular and more vigilant follow-ups are required for a substantial period to ensure that no local recurrence of upgradation is left unattended and untreated.

## Data Availability

All data produced in the present study are available upon reasonable request to the authors

## Author Declarations

I confirm that all relevant ethical guidelines have been followed and that any necessary approvals from IRB and/or ethics committee have been obtained.

## Highlights

- Intraductal Papilloma excision is important as although benign on the look, it has a high rate of upgradation to carcinoma.
- The classical surgical excision approach is the gold standard but it leaves a scar and has higher complication rates.
- Vacuum-assisted excision for Intraductal papilloma can be employed as a useful technique for better surgical results and fewer complications
- VAE provides better cosmetic results as well as less recurrence.
- Vigilant follow-up checks for a substantial period can be helpful in catching the recurrence and managing it.

